# Magnitude and causes of under-five mortality in the North Shewa Zone, Ethiopia

**DOI:** 10.1101/2022.11.15.22282351

**Authors:** Delayehu Bekele, Ruoqing R. Wang-Cendejas, Frederick G. B. Goddard, Bezawit M. Hunegnaw, Sebastien Haneuse, Tefera Bitye, Hemen Muleta, Yahya Mohammed, Mesfin Zeleke, Kimiko Van Wickle, Chalachew Bekele, Grace J. Chan

## Abstract

**Introduction:** Globally around 5 million children under the age of 5 years die every year. Ethiopia is one of five countries that contribute to more than half of these deaths. In the absence of a national vital registry system in Ethiopia, there is a lack of high-quality data regarding causes of death.

**Methods:** We conducted a verbal autopsy study among under-five deceased children at the Birhan health and demographic site (HDSS) located in the North Shewa zone, Ethiopia. All deaths of under five children, which occurred in the time period from March 1, 2019 to February 29, 2020 at the Birhan HDSS, were identified. The under-five mortality rate (U5MR) was computed using a Kaplan-Meier approach. The world health organization verbal autopsy questionnaire was administered to mothers or caregivers of the deceased child after a minimum mourning period of 42 days. The causes of death were then determined by the InterVA-5 algorithm.

**Results:** There were 90 under-five deaths resulting in an estimated U5MR of 45.3 per 1000 live births. The majority (62.2%) of deaths occurred during the neonatal period. The leading causes of death in under-five children were birth asphyxia (20, 27.0%), prematurity (19, 25.7%) and meningitis and encephalitis (8, 10.8%) while leading causes of neonatal deaths were birth asphyxia (19, 38.0%), prematurity (18, 36.0%) and neonatal sepsis (6, 12.0%).

**Conclusions:** There is a high under-five mortality rate at the study site, with most deaths occurring in the neonatal period. Birth asphyxia and prematurity contributed to more than half of the under-five deaths suggesting that follow-up and management during pregnancy, labor and delivery should be strengthened to avert a significant proportion of under-five deaths.

**What is already known on this topic**

- Ethiopia is one of the countries with the highest deaths of under 5 children but because absence of national vital registry system there is lack of high-quality data regarding causes of death.

**What this study adds**

- Birth asphyxia and prematurity are the commonest causes of death contributing to more than half of the under-five deaths.

**How this study might affect research, practice or policy**

- Follow-up and management during pregnancy, labor and delivery should be strengthened to avert a significant proportion of under-five deaths.

## Background

In 2020, an estimated 5 million children under the age of five years died, mostly from preventable and treatable causes (1). Sub-Saharan Africa and southern Asia account for more than 80% of these deaths, even though these regions only account for 53% of the global live births. Half of all under-5 deaths in 2020 occurred in just five countries: Nigeria, India, Pakistan, the Democratic Republic of the Congo and Ethiopia.

The Sustainable Development Goals aims to end preventable deaths of neonates and children under 5 years of age by 2030, with all countries aiming to reduce neonatal mortality to at least as low as 12 per 1,000 live births and U5MR to at least as low as 25 per 1,000 live births(2). Ethiopia’ s progress toward reducing the neonatal, infant, and under-five mortality in 2019 lies at 30, 43 and 55 deaths per 1000 live births, respectively(3).

Globally, infectious diseases, including pneumonia, diarrhea and malaria, along with preterm birth, birth asphyxia and trauma, and congenital anomalies remain the leading causes of death for children under five (4). However, there is a lack of high-quality data in Ethiopia to accurately describe and determine the causes of death among these children. This is in part because Ethiopia does not yet have a complete vital registration system.

Verbal Autopsy (VA) is a technique developed to ascertain the causes of death using a detailed questionnaire and is especially important in low resource settings where complete vital registration systems with medical certification of deaths and cause of death information is lacking (5). It obtains information on symptoms, signs, and other relevant events during the illness leading to death. The information is collected from caregivers of the deceased, most often the family members or others who have detailed knowledge of the overall circumstances, signs, and symptoms prior to death.

To date, attempts to determine the causes of death among under five children in Ethiopia using VA are scarce. In an early study done at the Butajira Health and Demographic Surveillance System (HDSS), southern Ethiopia, in 1987, the estimated cumulative under five mortality rate was 293 per 1000 live births and the infant (0-11 months old) mortality rate was 136 per 1000 live births. The major probable causes of death using VA was determined to be diarrhoeal disease or acute respiratory infections (6). At the Kilte-Awlaelo HDSS located in North Ethiopia, a longitudinal study covering the year 2009 - 2017 estimated an U5MR of 35.62 per 1000 live births, and that the top five leading causes of deaths using VA were bacterial sepsis, prematurity, intestinal infection disease/diarrheal disease, acute Lower Respiratory Infections, and birth asphyxia (7).

Apart from the previously mentioned studies, there are no other studies done using VA to estimate U5MR in Ethiopia and in the Amhara region in particular. Hence in this study, was done to determine the total number of deaths, mortality rates, and causes of deaths based on the verbal autopsy among neonates (0-27 days), infants (<1 year), and under-five children (<5 years) at the Birhan HDSS located in the North Shewa zone, Ethiopia.

## Methods

### Study design and period

The Birhan demographic and health surveillance system follows a population-based longitudinal study design (8, 9). This mortality study is nested within the Birhan HDSS and includes the time period from March 1, 2019 and February 29, 2020.

### Study setting and population

The study was conducted at the Birhan HDSS site which is located in the North Shewa zone of the Amhara regional state of Ethiopia (Figure 1)(9). The Birhan field site serves as a platform for community and facility-based research with a focus on maternal and child health. The catchment area covers 16 Kebeles (the lowest administration structures) across two districts namely Angolela-Tera a predominantly highland and Shewarobit-Kewot which is predominantly low land. The population was predominantly (82.6%) rural(9).

**Figure 1:**
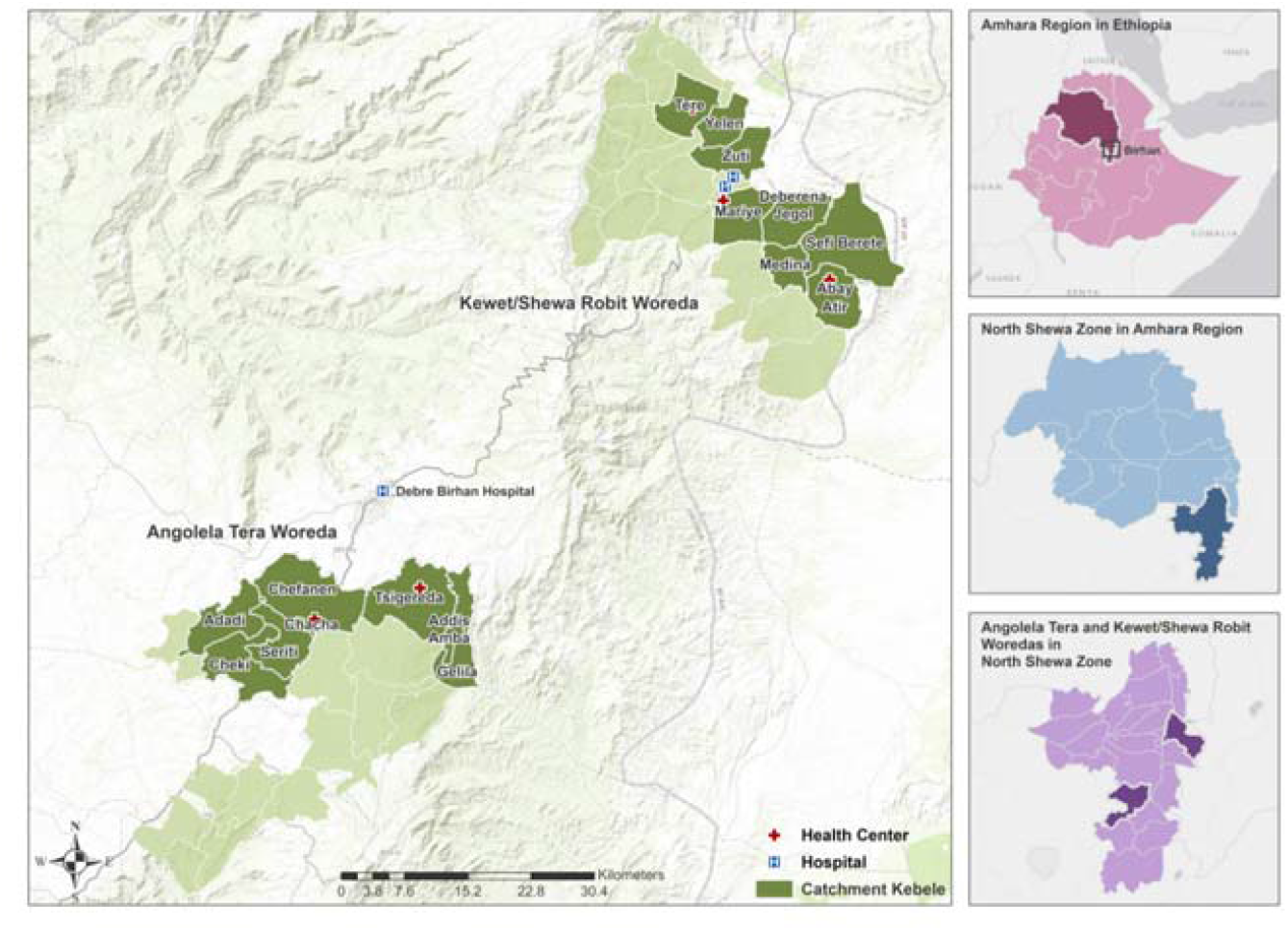
BIRHAN for Mothers and Children Field Sites.

There were 84,737 individuals residing in the study area during the study period. Out of which, 11,120 under-five children enrolled in the study during the study period were the source population for this study.

### Data collection tools and procedure

The 2016 WHO 2016 verbal autopsy (VA) instrument was used (10). The VA questionnaire was translated into the local language Amharic and uploaded to tablets on open data kit (ODK) platform. A pilot test was conducted to assure clarity of the questions, local perceptions of specific symptoms and signs, and accuracy and completeness of questionnaires. The pilot test was also used to estimate the time needed to complete the questionnaire. The way the questions were phrased was modified based on the feedback from the pilot.

The list of all mortality cases recorded during the study period in the Birhan HDSS and its pregnancy and birth cohort was provided to the VA team. After a minimum recommended mourning period (42 days) has passed, the VA questionnaire was administered to mothers or caregivers of the deceased child. The VA interview was conducted by trained data collectors who are nurses and midwives by background. The data collectors were trained for ten days on the data collection tool, purpose of study, and process of data collection. To maintain the quality of the data; the study team performed intensive field supervision with direct observation of the data collection in the field and provided feedback and regular refresher trainings.

### Data management and analysis

All data analysis was conducted using STATA 17 and frequencies and cross-tabulations were used to summarize descriptive statistics. The U5MR was computed using a Kaplan-Meier (KM) approach. Specifically, the KM method calculates the risk of surviving at each age (in days) leading up to age 5 within 1 year of observation. To this end, the KM method first calculates the conditional probability of survival at each age (in days) and multiplies the individual probabilities of survival, resulting in a cumulative probability of survival. The probability of death (or cumulative mortality rate) is subtracting the cumulative probability of survival from 1. For the conditional probability of survival at each age, it is calculated as a ratio between the number of survivors and number of participants at risk.

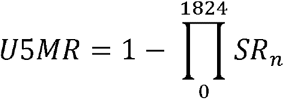

where *SR*_*n*_ stands for the conditional probability of survival at an age (in days) between March 1, 2019 and February 29, 2020.

The cause of death was ascertained by InterVA-5 software model (11). The InterVA algorithm was developed based on Bayes’ rule for conditional probabilities (14). Based on demographic indicators and reported symptoms collected in a VA interview and a set of evidence-based and physician-derived conditional probabilities of the likelihood of each indicator and symptom, the algorithm produced the propensities for each possible cause (14). For a set of deaths, population-level fraction of mortality from each cause was computed by the sum of the largest propensities for each cause. For each death, the top three causes were determined by the largest propensities, if they fall above a threshold; otherwise, the cause is “indeterminate” (14).

### Ethics approval

Ethical approval was obtained for the study from the Saint Paul’ s Hospital Millennium Medical College and the Harvard T. H. Chan School of Public Health. All the heads of family or involved people for the interview for the verbal autopsy gave written informed consent. All the information obtained from the interview were kept confidential and are not shared with a third party.

### Patient and public involvement

Patients and/or the general public were not involved in the initial design of this study but a community advisory board was established and involved during the execution of this study.

## Results

There were a total of 447 deaths in the Birhan HDSS between March 1, 2019 and February 29, 2020, among which 90 (20.1%) were under-five deaths. Out of the 90 under-five deaths, 67.8% were males and 74.4% were from rural areas (Table 1). 56 (62.2%) were neonatal deaths under 28 days, and 80 (88.9%) were infant deaths under 1 year. There were 33 early neonatal deaths (0-6 days), 23 late neonatal deaths (7-27 days), 24 postneonatal deaths (28 days-<1 year), and 10 child deaths between the ages of 1 and 5 years.

**Table 1:** Distribution of deceased child by age, gender and place of residence among under 5 children (<5 years) in the North Shewa zone, Amhara Region, Ethiopia, March 2019 - February 2020

|  | N | n (%) |
| --- | --- | --- |
| **Age group** |  |  |
| Under-five (< 5 years) | 90 |  |
| Newborns (0-27 days) | 90 | 56 (62.2) |
| Postneonatal (28 days - <1 year) | 90 | 24 (26.7) |
| Children (1 - <5 years) | 90 | 10 (11.1) |
| **Neonatal age group** |  |  |
| Early Neonates (0-6 days) | 56 | 33 (58.9) |
| Late Neonates (7-27 days) | 56 | 23 (41.1) |
| **Sex** |  |  |
| Male | 90 | 61 (67.8) |
| Female | 90 | 29 (32.2) |
| **Woreda** |  |  |
| Angolela Tera | 90 | 53 (58.9) |
| Kewet Shewarobit | 90 | 37 (41.1) |
| **Residence** |  |  |
| Rural | 90 | 67 (74.4%) |
| Urban | 90 | 23 (25.6%) |

As shown in Table 2, the total number of male deaths was higher than the number of female deaths and this is true for all age subgroups. But the gender difference in deaths at different ages were not statistically significant (Chi-square, χ²=1.115, p=0.774).

**Table 2:**
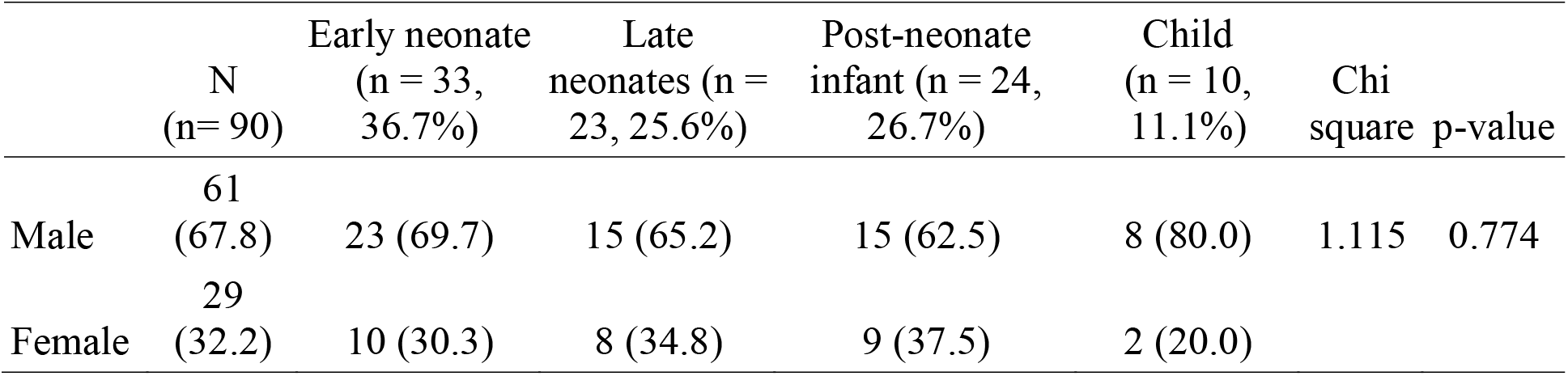
Age-sex distribution among deceased under 5 children (<5 years) in the North Shewa zone, Amhara Region, Ethiopia, March 2019 - February 2020

Figure 2 shows the Kaplan-Meier survival function and 95% confidence interval and Table 3 shows the results of the Kaplan-Meier analysis. The cumulative probability of survival at the end of the neonatal period is 0.9720 (95% CI: [0.9638, 0.9784]), resulting in a neonatal mortality rate of (1-0.9720)*1,000 = 28.0 per 1,000 live births. For infants (<1 year), the probability of survival is 0.9600 (95% CI: [0.9504, 0.9677]) with a mortality rate of 40.0 per 1,000 live births, and for children under 5, the probability of survival is 0.9547 (95% CI: [0.9446, 0.9630]) with a mortality rate of 45.3 per 1,000 live births.

**Table 3:**
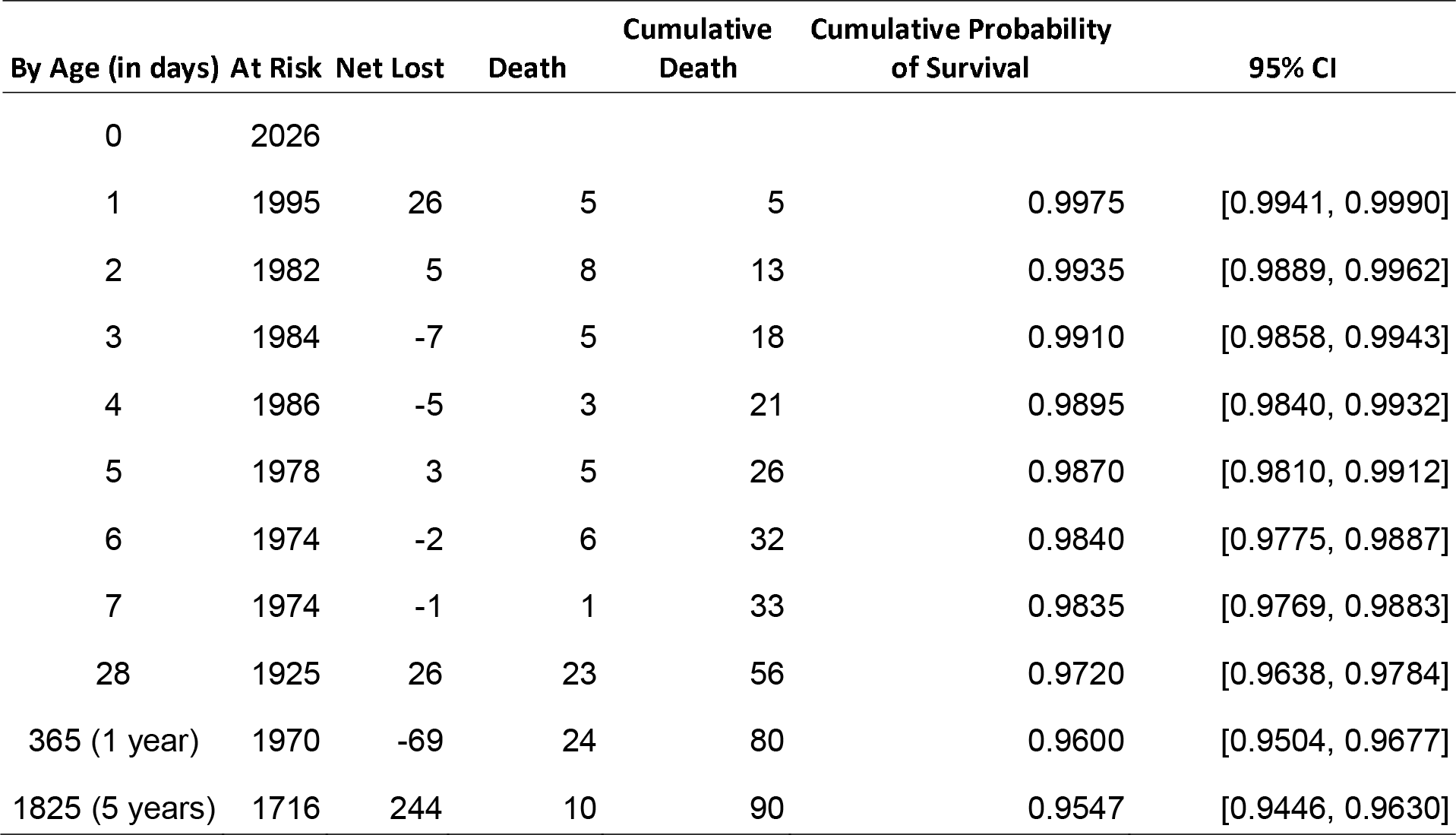
Kaplan-Meier Probability of Mortality and Probability of Survival by Age Group, March 2019 - February 2020

**Figure 2:**
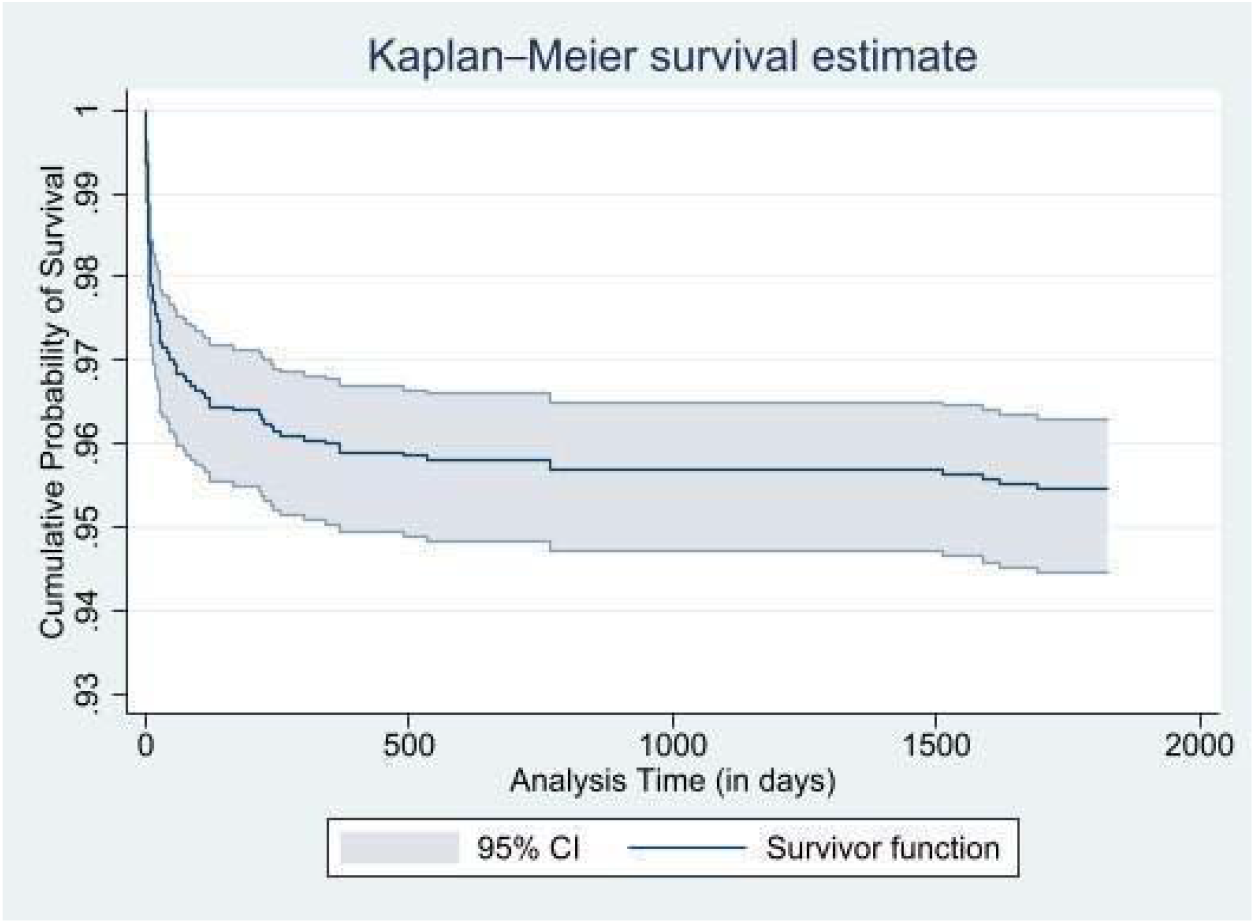
Kaplan-Meier Survival Function.

Within the neonatal group, early neonates (0-6 days) had a mortality rate of 16.5 per 1000 live births, and the late neonates (7-27 days) had a figure of 11.5 per 1000 live births. For postneonatal infants (28 days - <1 year) and children 1-5 years, their mortality rates are 12.0 and 5.3, both per 1000 live births. The death rate for the different age groups are shown in Table 4.

**Table 4:**
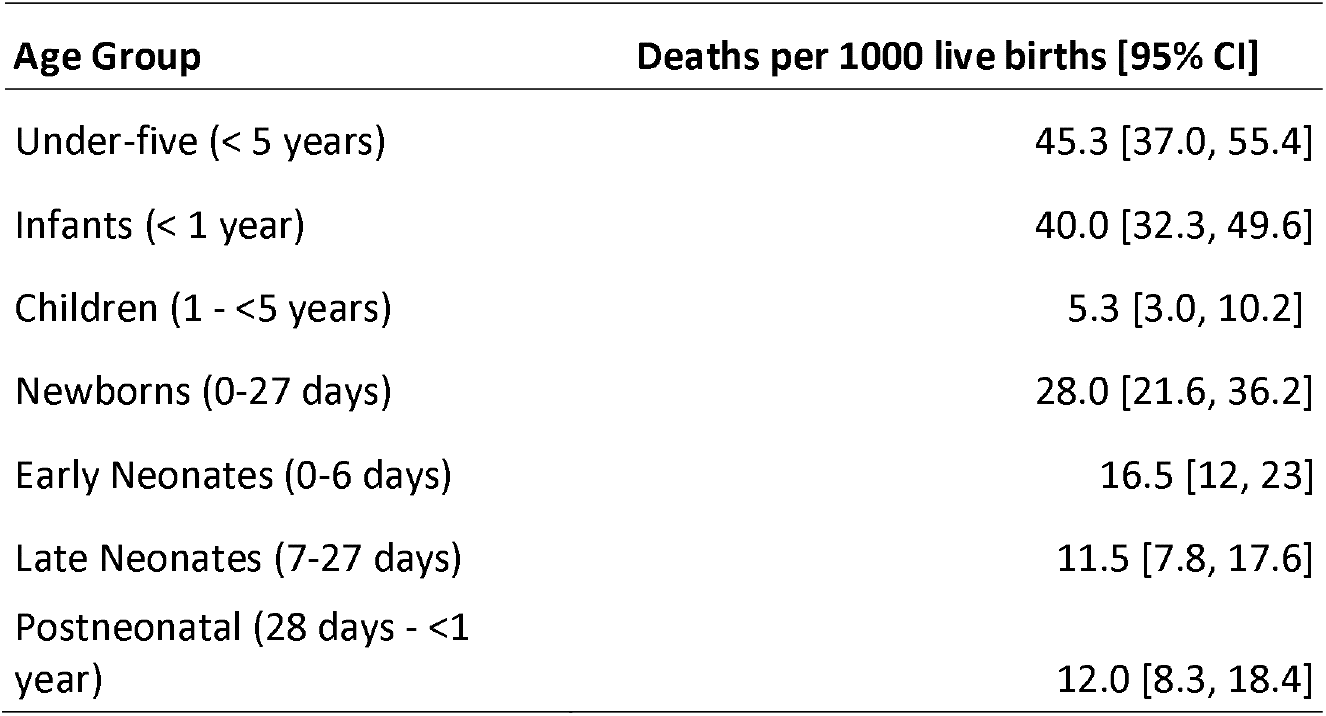
Mortality rates among under 5 children (<5 years) and various age subcategories in the North Shewa zone, Amhara Region, Ethiopia, March 2019 - February 2020

| Age Group | Deaths per 1000 live births [95% CI] |
| --- | --- |
| Under-five (< 5 years) | 45.3 [37.0, 55.4] |
| Infants (< 1 year) | 40.0 [32.3, 49.6] |
| Children (1 - <5 years) | 5.3 [3.0, 10.2] |
| Newborns (0-27 days) | 28.0 [21.6, 36.2] |
| Early Neonates (0-6 days) | 16.5 [12, 23] |
| Late Neonates (7-27 days) | 11.5 [7.8, 17.6] |
| Postneonatal (28 days - <1 year) | 12.0 [8.3, 18.4] |

### Causes of death

Out of the 90 under-five mortality cases, 74 (82.2%) cases have a cause of death determined by InterVA-5. The remaining 17.3% had indeterminate cause of death due to inadequate information or missing critical information. The most common causes of death in under-five children were birth asphyxia (20, 27.0%), prematurity (19, 25.7%) and meningitis and encephalitis (8, 10.8%). The major causes responsible for female under-five mortality were birth asphyxia (9, 34.6%), neonatal sepsis (4, 15.4%) and meningitis and encephalitis (3, 11.5%). The leading causes of death among male children were prematurity (17, 35.4%), birth asphyxia (11, 22.9%) and meningitis and encephalitis (5, 10.4%). These results are shown in Table 5.

**Table 5.**
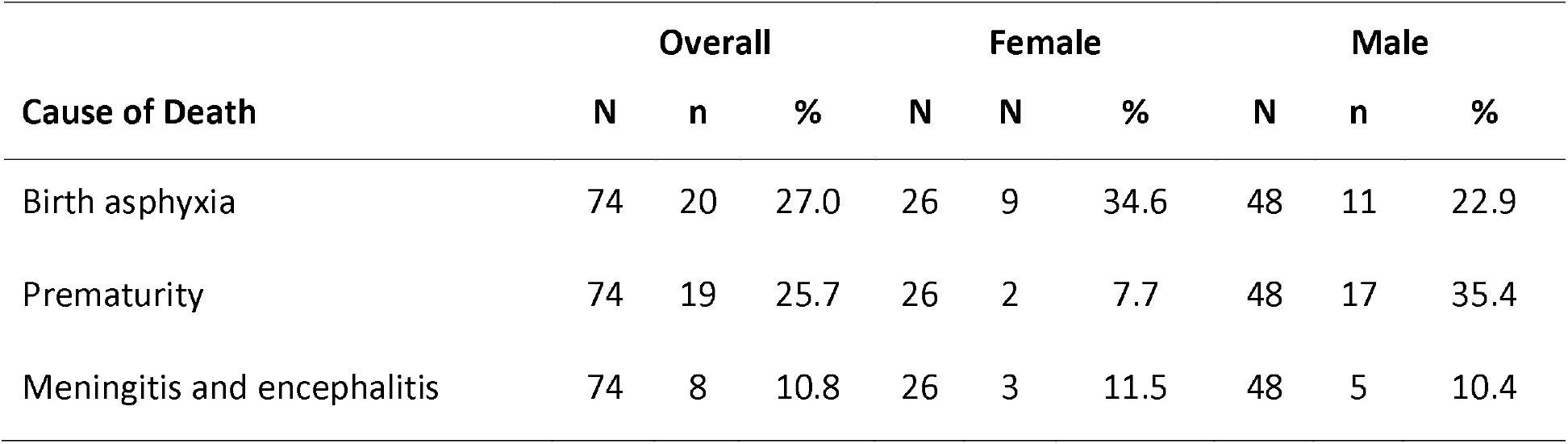
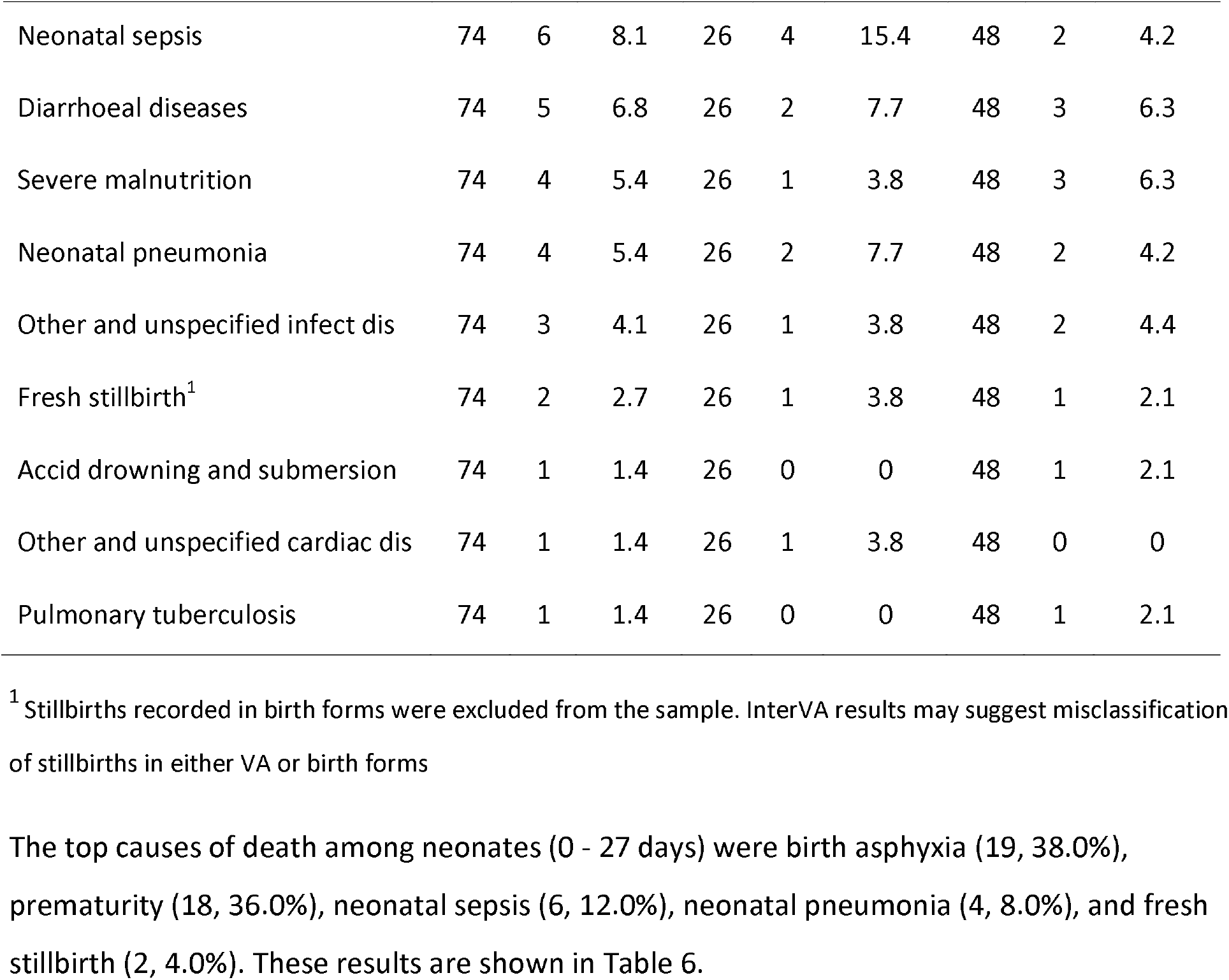
Causes of Death among under-five children (<5 years) in the North Shewa zone, Amhara Region, Ethiopia, March 2019 - February 2020

The top causes of death among neonates (0 - 27 days) were birth asphyxia (19, 38.0%), prematurity (18, 36.0%), neonatal sepsis (6, 12.0%), neonatal pneumonia (4, 8.0%), and fresh stillbirth (2, 4.0%). These results are shown in Table 6.

**Table 6:**
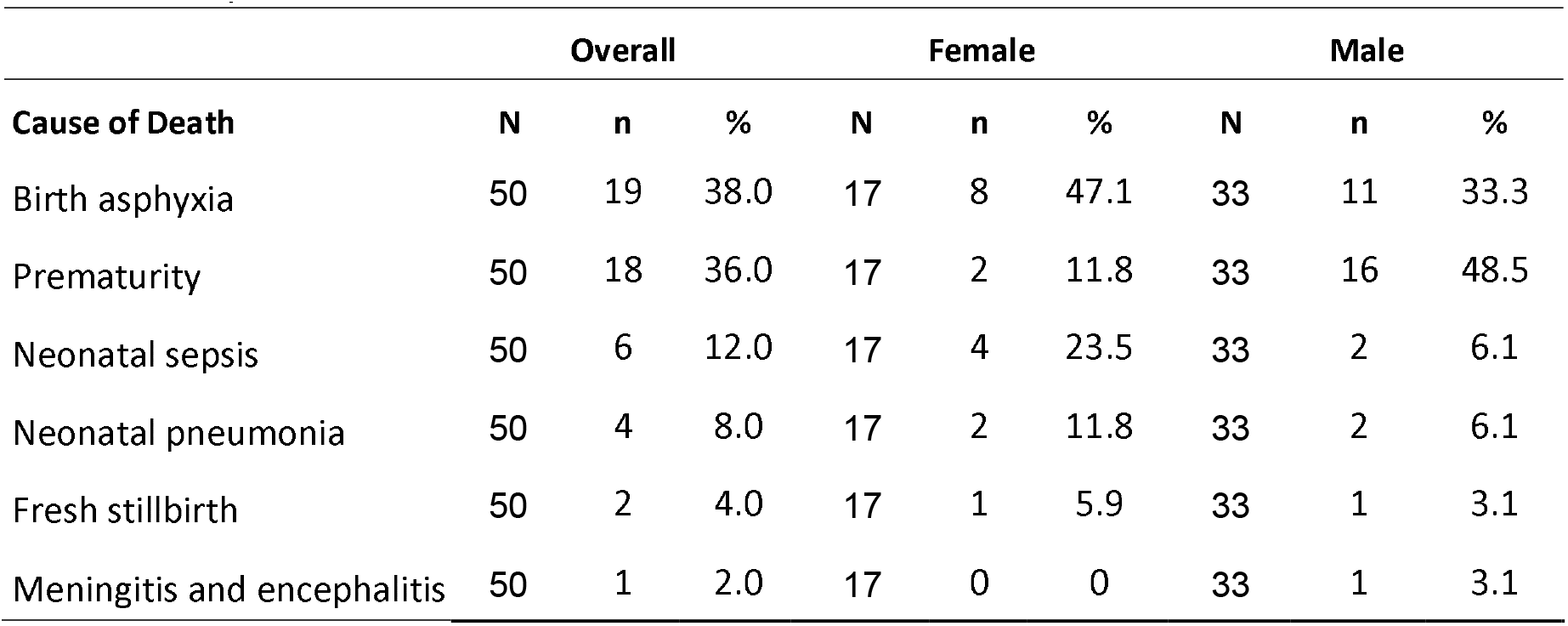
Causes of Death among neonates (0-27 days) in the North Shewa zone, Amhara Region, Ethiopia, March 2019 - February 2020

| Cause of Death | Overall |  |  | Female |  |  | Male |  |  |
| --- | --- | --- | --- | --- | --- | --- | --- | --- | --- |
|  | N | n | % | N | n | % | N | n | % |
| Birth asphyxia | 50 | 19 | 38.0 | 17 | 8 | 47.1 | 33 | 11 | 33.3 |
| Prematurity | 50 | 18 | 36.0 | 17 | 2 | 11.8 | 33 | 16 | 48.5 |
| Neonatal sepsis | 50 | 6 | 12.0 | 17 | 4 | 23.5 | 33 | 2 | 6.1 |
| Neonatal pneumonia | 50 | 4 | 8.0 | 17 | 2 | 11.8 | 33 | 2 | 6.1 |
| Fresh stillbirth | 50 | 2 | 4.0 | 17 | 1 | 5.9 | 33 | 1 | 3.1 |
| Meningitis and encephalitis | 50 | 1 | 2.0 | 17 | 0 | 0 | 33 | 1 | 3.1 |

Table 7 shows that birth asphyxia (20, 29.4%), prematurity (19, 27.9%), meningitis and encephalitis (7, 10.3%), neonatal sepsis (6, 8.8%), and diarrhoeal diseases (5, 7.4%) are the top causes of deaths for infants (< 1 year).

**Table 7.**
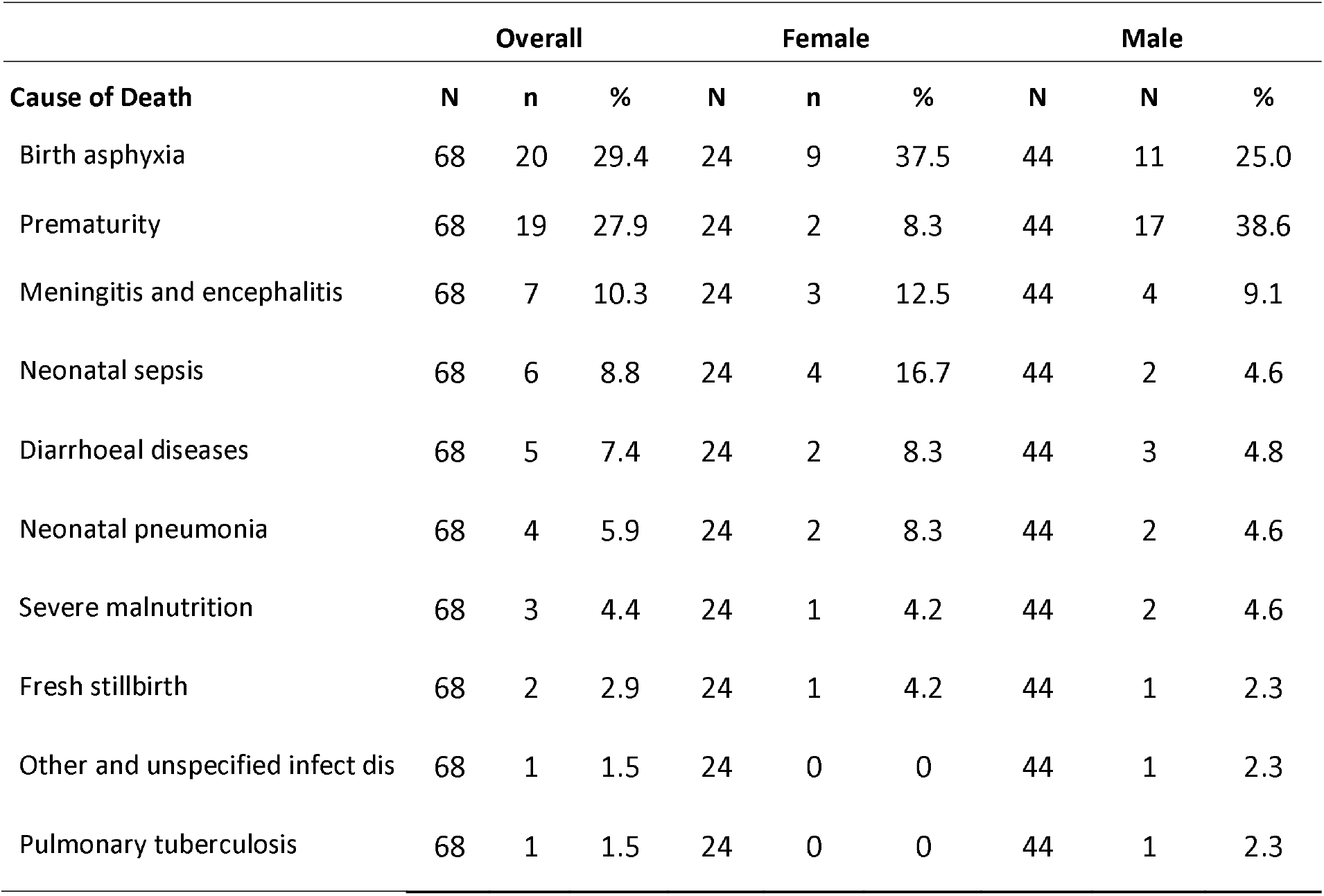
Causes of Death among infants (< 1 year) in the North Shewa zone, Amhara Region, Ethiopia, March 2019 - February 2020

## Discussions

Our study demonstrated the magnitude and common causes of deaths among under five children, neonates, and infants. The latest data from the 2019 Ethiopia Mini DHS showed that the national and the Amhara regional U5MR were 59 and 69 deaths per 1,000 live births respectively. In our study, the U5MR was 45.3 per 1,000 live births in two woredas in Amhara. The national neonatal and infant mortality rates were 33 and 47 per 1,000 live births respectively while these numbers were 28.0 and 40.0 per 1,000 live births respectively in our current study. The mortality rates in our study are numerically lower than the national and regional estimates, but it is difficult to make direct comparisons because of the methodological differences in analysis. The EDHS mortality estimates implemented a survey method based on a probability (expressed as a rate per 1,000 live births) of a child born in a specific year or period dying before reaching the age of five years. The national and the regional U5MRs are also calculated for the 5-year and 10-year period, respectively, before the survey to ensure that there are sufficient cases to produce statistically reliable estimates. Our current study is based on a one-year observation in two woredas in the Amhara region and implements a Kaplan-Meier approach to calculate the probability of surviving at each age (in days) leading up to age 5 by design. This difference in the methodological approaches may partially explain the difference in results.

There is a significant difference in the gender contribution among the deceased children with more than two thirds (67.8%) being male. A similar study from the Kilite-Awlaelo HDSS in the northern part of Ethiopia has also shown the same trend, even though the gender gap was not as wide as the one in the current study, and males have contributed to 55.5% of all under-five deaths (7). Other studies from and outside Ethiopia haven’ t shown a similar trend(12). A detailed analysis of the sex differentials among the mortalities is being dealt in a separate paper.

62.2% of deaths occurred in the first 28 days of life while the postneonatal infant (28 days - <1 year) and child (1 year - <5 years) deaths contribute 26.7% and 11.1% of deaths. The contribution of neonatal mortality is much higher than previous studies (13). To investigate possible under-ascertainment of mortality in the postneonatal infant (28 days - <1 year) and child (1 year - <5 years) age groups, we performed sensitivity analyses to estimate a range of possible mortality rates for postneonatal infants (28 days - <1 year), children (1 year - <5 years), and under-five children. We estimated mortality rates by considering changes in the number of postneonatal infant deaths (28 days - <1 year), child deaths (1 year - <5 years) and both postneonatal infant and child deaths to reflect under-ascertainment between 5% and 20% in the data.

The results of sensitivity analyses indicated that if the under-ascertainment of postneonatal infant deaths was between 5% and 20%, the U5MR ranged from 46 per 1000 live births to 48 per 1000 live births, respectively. If the under-ascertainment of child deaths was between 5% and 20%, the U5MR ranged from 46 per 1000 live births to 47 per 1000 live births, respectively. If both the post-neonates and child between 1 year and 5 years were under-ascertained by 5% to 20%, the U5MR ranged from 46 per 1000 live births to 50 per 1000 live births, respectively. These are shown in the supplemental document.

The top under-five causes of death from the current study include birth asphyxia, prematurity, meningitis and encephalitis, neonatal sepsis, and diarrheal diseases. They are different from the ones found in a meta-analysis, which reviewed the under-five causes of death in Ethiopia, 1990 - 2016. The leading under five causes of death in the study included acute lower respiratory infection (including pneumonia), severe acute malnutrition, sepsis, and diarrheal diseases(14). A more recent study in the Kilite-Awlaelo HDSS in Ethiopia found that bacterial sepsis, prematurity, intestinal infection disease, acute lower respiratory infections, and birth asphyxia were the major causes of under-five mortality, 2009-2017, based on the VA method (physician review but not the InterVA algorithm) (7).

The difference to national and regional results may be explained by the composition of the sample as well. Because of the distribution of the sample (high percentage of neonates and infants), the under-five causes of deaths are largely driven by neonatal and infant causes of deaths between March 2019 and February 2020.

The limitation of our study would be that the U5MR estimates are based on data of one-year between March 2019 and February 2020. The available data does not permit a survey method often used in nationally representative studies to calculate the probability (expressed as a rate per 1,000 live births) of a child born in a specific year or period dying before reaching the age of five years during a standard, 5- or 10-year period prior to survey.

The InterVA algorithm may not conclusively determine the causes of death for all the under-five mortality cases in the study. The causes of death are indeterminate in 17.8% which is similar to other similar studies. A study in Kenya done among under five children to determine the VA causes of death 15.7% of cases has an indeterminate cause of death (15). There is no process for cause of death attribution that leads to an absolute “truth” for every case. For this reason, the COD results are limited to the cases with determined causes. Moreover, this study is largely descriptive. Future analysis may consider additional statistical tests to determine any possible analytical inferences.

## Conclusion

U5MR is high with the majority occurring in the neonatal period. Birth asphyxia and prematurity which are related to the care received during pregnancy and peripartum period contributed for more than half of the under-five deaths. This suggests that proper follow-up and management during pregnancy, labor and delivery could prevent a significant proportion of under-five deaths.

## Data Availability

All data produced in the present study are available upon reasonable request to the authors

## Reference

1. World Health Organization. Child mortality (under 5 years): World Health Organization,; 2022 [Available from: https://www.who.int/news-room/fact-sheets/detail/levels-and-trends-in-child-under-5-mortality-in-2020.

2. United Nation. Transforming our world: The 2030 agends for sustainable development [Available from: https://sustainabledevelopment.un.org/content/documents/21252030%20Agenda%20for%20Sustainable%20Development%20web.pdf.

3. EDHS. Ethiopia Mini Demographic and Health Survey 2019: Key Indicators. Rockville, Maryland, USA: EPHI and ICF: Ethiopian Public Health Institute (EPHI) [Ethiopia] and ICF; 2019.

4. Liu L, Oza S, Hogan D, Chu Y, Perin J, Zhu J, et al. Global, regional, and national causes of under-5 mortality in 2000-15: an updated systematic analysis with implications for the Sustainable Development Goals. Lancet. 2016;388(10063):3027–35.

5. Thomas LM, D’ Ambruoso L, Balabanova D. Verbal autopsy in health policy and systems: a literature review. BMJ Glob Health. 2018;3(2):e000639.

6. Lozano R, Freeman MK, James SL, Campbell B, Lopez AD, Flaxman AD, et al. Performance of InterVA for assigning causes of death to verbal autopsies: multisite validation study using clinical diagnostic gold standards. Popul Health Metr. 2011;9:50.

7. Abraha HE, Belachew AB, Ebrahim MM, Tequare MH, Adhana MT, Assefa NE. Magnitude, trend, and causes of under-five mortality from Kilite-Awlaelo health demographic surveillance database, northern Ethiopia, 2009-2017. BMC Public Health. 2020;20(1):1465.

8. Chan GJ, Hunegnaw BM, Van Wickle K, Mohammed Y, Hunegnaw M, Bekele C, et al. Birhan maternal and child health cohort: a study protocol. BMJ Open. 2021;11(9):e049692.

9. Bekele D, Hunegnaw BM, Bekele C, Van Wickle K, Tadesse F, Goddard FGB, et al. Cohort Profile: The Birhan Health and Demographic Surveillance System. Int J Epidemiol. 2021.

10. Nichols EK, Byass P, Chandramohan D, Clark SJ, Flaxman AD, Jakob R, et al. The WHO 2016 verbal autopsy instrument: An international standard suitable for automated analysis by InterVA, InSilicoVA, and Tariff 2.0. PLoS Med. 2018;15(1):e1002486.

11. Byass P, Hussain-Alkhateeb L, D’ Ambruoso L, Clark S, Davies J, Fottrell E, et al. An integrated approach to processing WHO-2016 verbal autopsy data: the InterVA-5 model. BMC Med. 2019;17(1):102.

12. Ramy Mohamed Ghazy MMF, Abdel-Rahman Omran, Mohamed Mostafa Tahoun. Causes of under-five mortality using verbal autopsy and social autopsy studies (VASA) in Alexandria, Egypt, 2019. Journal of Global Health Reports. 2020; 4.

13. Sushil Dalal KP, Md Abu Bashar. Study of Death among Children Below Five Years of Age and its Relation with Socio Economic Status and Place of Residence Using Verbal Autopsy as a Tool in Deharadun. Indian Journal of Public Health Research & Development,. January 2020,;Vol. 11, No. 01.

14. Wubegzier Mekonnen NA, Wubetsh Asnake, Zekarias Sahile3, Damen Hailemariam1. Under five causes of death in Ethiopia between 1990 and 2016: Systematic review with meta-analysis. Ethiop J Health Dev. 2020;Vol. 34 (2).

15. Oti SO, Kyobutungi C. Verbal autopsy interpretation: a comparative analysis of the InterVA model versus physician review in determining causes of death in the Nairobi DSS. Popul Health Metr. 2010;8:21.

